# Abdominal Myosteatosis and Cognitive Decline in a Biracial Cohort: Insights from CARDIA Study

**DOI:** 10.1101/2025.01.17.25320741

**Authors:** Adrianna Acevedo-Fontánez, Caterina Rosano, Kristine Yaffe, J Jeffrey Carr, James G. Terry, Sangeeta Nair, Emma Barinas-Mitchell, Ryan K. Cvejkus, Iva Miljkovic

**Author notes:** **CORRESPONDING AUTHOR**: Iva Miljkovic, MD, PhD, Department of Epidemiology, Graduate School of Public Health, University of Pittsburgh, Pittsburgh, PA; 130 De Soto Street, Room A524 Public Health, Pittsburgh, PA 15261.

## Abstract

**Objective:** Skeletal muscle fat infiltration (myosteatosis) increases with age and is an emerging risk factor for dementia. We aimed to determine the association between myosteatosis and cognitive decline among middle-aged White and Black Americans.

**Methods:** Data were on men (n=1,080; 41.9% Black) and women (n=1,432; 49.0% Black) from the CARDIA study. CT-measured abdominal intermuscular fat (IMAT) volume was assessed at baseline. Cognition was assessed by Digit Symbol Substitution (DSST), Rey Auditory Verbal Learning (RAVLT), and Stroop Test at baseline and 5-year follow-up. Multivariable linear regression was used to assess associations of IMAT with cognitive change.

**Results:** Participants were aged 50.2 (3.6) years and had IMAT of 2.3 (1.6) cm^3^, 5-year change in DSST of -2.8% (21.8), RAVLT 2.8% (17.5) and Stroop 6.5% (49.5). Greater IMAT was associated with steeper DSST decline (β =-0.52 points per SD, p-value=0.035), but not with Stroop or RAVLT. Stratified by race, greater IMAT predicted DSST decline among White (β =-0.73, p =0.044), but not Black (β =-0.44, p =0.195), participants.

**Conclusions:** Abdominal myosteatosis may be a novel risk factor for decline in psychomotor speed, especially in middle-aged Whites. Further research on mechanisms, including metabolic mediators, is warranted to understand myosteatosis’s role in mid-life cognitive decline.

## INTRODUCTION

Cognitive functioning involves the ability to perceive and react, process and understand, make decisions, and produce appropriate responses to the environment. Although the natural aging process can lead to some cognitive changes, there is growing evidence that certain types of cognitive decline are recognizable as early signs of a neurodegenerative disorder, which could eventually result in dementia.^1^ Alzheimer’s disease is the most common form of dementia, affecting around 6.5 million people aged 65 and above in the United States.^2^ In the US, 22% of individuals 65 years and older are living with mild cognitive impairment and 10% with dementia.^3^ Further, Alzheimer’s Disease and related dementias (ADRD) prevalence is higher in women (12.2%) than men (8.6%).^4^ ADRD disproportionally affects persons of African Ancestry^4–6^, and those with lower education^3,7^, and there is an urgent need to delay or prevent the onset of disease to promote years free from disability in this rapidly growing demographic.

Midlife is a critical period for exposure to cardiometabolic and other risk factors that could cause subtle changes leading to later-in-life diseases. A high BMI during midlife is linked to an increased risk of developing dementia, a concern heightened by the global obesity epidemic.^8–11^ Indeed, increased adiposity, in particular, ectopic adiposity, has been shown to be associated with and a predictor for impaired cognitive performance, accelerated cognitive decline, and dementia.^12^ Ectopic adiposity is a phenomenon seen during weight gain and aging,^13–16^ when adipocytes (cells specialized in fat storage) reach their storage capacity. Fat then begins building in surrounding (non-adipose tissue) organs, such as the skeletal muscle, and this fat infiltration into the muscle is known as myosteatosis.^13^ Myosteatosis is an understudied ectopic adiposity depot of potentially high importance in People of African Ancestry, given their greater levels of myosteatosis^17^ as compared to White individuals. Furthermore, it has been shown that inter-muscular adipose tissue (IMAT) can impair the normal physiologic function of the muscle, making it a physiologically relevant adiposity depot that may play a key role in the risk of metabolic abnormalities such as insulin resistance and type 2 diabetes,^16,18,19^ which are also linked to dementia.

To date, just a few studies have assessed the specific relationship of myosteatosis in the lower extremities and cognitive function, mainly in cross-sectional studies or studies of older age adults. Higher myosteatosis in the calf and thigh has been associated with lower information processing speed^20^ and with poorer psychomotor function and visual learning.^21^ Myosteatosis was associated with an increased rate of brain aging measured through an MRI-assessed brain age gap (discordance between actual biological brain age and a prediction of brain age).^22^ More recently, a study found an independent association between increasing thigh myosteatosis and cognitive decline.^23^ It remains unclear if abdominal myosteatosis has the same associations with cognition and aging-related cognitive decline, and if these associations might be relevant in middle-aged Black and White men and women and are independent of cardiometabolic conditions. Studying the impact of adipose tissue accumulation in the muscle on cognitive decline among individuals of different ethnicities and sexes could help identify new pathways associated with dementia and improve our understanding of racial disparities. Thus, we examined the association between abdominal IMAT and cognitive function decline in a representative cohort of middle-aged White and Black American men and women. We aimed to confirm the findings of previous reports on the cross-sectional association between IMAT and DSST. We also hypothesized that participants who have higher abdominal IMAT will have a greater decline over time in cognitive function as measured by DSST, the Rey Auditory Verbal Learning Test (RAVLT)^24^, and the Stroop Test, which capture processing speed, memory, and task shifting, respectively. We further hypothesize that associations will be independent of sex, age, adiposity-related conditions (obesity, diabetes, hypertension, lifestyle risk factors), and education.

## METHODS

### Population

The Coronary Artery Risk Development in Young Adults (CARDIA) study began in 1985 with the recruitment of 5,115 participants aged 18 to 30 years at field centers located in Birmingham, AL, Chicago, IL, Minneapolis, MN, and Oakland, CA.^25^ Recruitment was balanced for equal distribution of Black and White female and male participants, age (18–24, 25–30 years), and education (≤12 years, >12 years). The current study includes data from participants who agreed to undergo an abdominal CT scan at the year 25 (Y25) examination. A total of 3,499 participants were examined in the clinic at Y25, representing 68.4% of the original cohort. Out of these 3,499 patients, 3,172 had abdominal CTs; 327 were eliminated due to weight, not being able to fit in the CT scanner, or pregnancy. Of the 3,172 participants who had abdominal CT scans, a total of 2,512 (79.2%) had complete details of abdominal fat and muscle composition, Digit Symbol Substitution Test (DSST), and other key factors such as age, sex, race, education, BMI, systolic blood pressure, diastolic blood pressure, lipid-lowering medication use, smoking status, alcohol consumption, and physical activity.

The proposal for the present study was approved by the CARDIA Publications and Presentations Committee. All participants provided written informed consent, and institutional review boards from each field center and the coordinating center approved the study annually.

### CT measures of Adiposity

Participants underwent multidetector CT chest and abdomen scans using a standardized protocol.^26,27^ The scans were performed at CARDIA field centers using 64-channel multidetector GE CT scanners (GE Healthcare Milwaukee, WI) at the Birmingham, AL, and Oakland, CA, centers and Siemens CT scanners (Siemens, Erlangen, Germany) at the Chicago, IL, and Minneapolis, MN, centers. CT images were electronically transmitted to the central CT reading center located at Wake Forest University School of Medicine, Winston-Salem, NC. Adipose tissue depots were measured volumetrically within a 10-mm block of 10 × 1 mm or 8 × 1.25 mm contiguous slices based on the nominal slice thickness produced by the specific scanner centered at the level of the disk between the 4th and 5th lumbar vertebrae as previously described.^27,28^ Medical Image Processing, Analysis, and Visualization (MIPAV) software was used to quantify subcutaneous (SAT) and VAT volume.

Abdominal muscle composition (lean, IMAT, and total) was measured from CT images covering the lower abdomen, which included four paired muscle groups: psoas, paraspinous, lateral oblique and rectus abdominis. ^27,28^ The abdominal muscles were measured at the L3–L4 level to avoid changes in muscle orientation related to the pelvic bones in some individuals at the L4–L5 level.

Each muscle boundary was manually traced. Tissues within muscle with attenuation between −190 and −30 Hounsfield units (HU) were defined as adipose tissue, and those with attenuation between −29 and 160 HU as lean tissue. Measures of left and right muscles in each group were highly correlated, so mean adipose, lean, and total volumes of the left and right sides were calculated and analyzed for all abdominal muscles. Analysis reliability of CT measures was assessed through blinded intra- and inter-reader re-reads of 158 scan pairs (approximately 5%). Overall (intra- and inter-reader) technical error in the re-analysis of 158 pairs of scans was 6.0% for VAT and 7.7% for psoas muscle total volume with correlations for re-reads >0.95 for each measure.

### Cognitive Function Tests

A battery of three standardized tests to measure cognitive function was administered at the Year 25 and Year 30 examinations. The Digit Symbol Substitution Test (DSST)^29^ is a test of information processing speed with scores ranging from 0 to 133, with a higher score indicating better cognitive performance. The DSST was administered by trained staff. Participants were asked to do a 10-symbol practice to assess their understanding of the test where the interviewer would fill the first 3 boxes of the sample section and then ask the participants to fill the remaining 7 boxes. Each participant had 90 seconds to complete as many DSST boxes as possible, going from left to right in order and without skipping any box. The number of correct responses was summed and scored as correct if they were clearly identifiable as the keyed symbol, even if it was drawn imperfectly (“v” instead of “u”) or if they were corrections of an incorrect symbol. The absolute change in the 5-year change in DSST score was calculated by subtracting the total DSST score in year 25 from the total DSST score in year 30.

The CARDIA Study also administered two additional cognitive tests: Rey Auditory Verbal Learning Test (RAVLT),^24^ measuring episodic memory, and the Stroop Color and Word Test (SCWT),^30^ which assesses the ability to inhibit cognitive interference, which occurs when the processing of a stimulus feature affects the simultaneous processing of another attribute of the same stimulus. The RAVLT assesses the ability to memorize and retrieve words (verbal memory) after several presentations of the word list immediately one after another, and then after a delay of 10 min.^24^ The range of scores on the delayed test is 0–15, with higher scores indicating better performance. The Stroop includes three subtests that evaluate the ability to view complex visual stimuli and to respond to one stimulus dimension while suppressing the response to the other dimensions.^30^ Each subtest is scored by summing the number of errors and the time required for completion. An interference score is calculated by subtracting the score on the incongruent subtest from the congruent subtest. Stroop interference scores can be negative or positive, and a higher interference score indicates worse performance on the task.

### Lifestyle and Comorbidities

At the baseline visit, participants reported age, sex, and years of education completed. Clinic visit procedures were standardized and consistent across examinations, as previously published in detail.^25^ Blood pressure was measured in triplicate after a 5-minute rest using an automated blood pressure monitor (Omron model HEM907XL; Omron Healthcare Inc., Lake Forest, IL). The average of the 2nd and 3rd readings was used to determine mean systolic blood pressure (SBP) and diastolic blood pressure (DBP). Hypertension was defined using the American College of Cardiology (ACC) (Whelton et al., 2017) definition as SBP ≥ 130, DBP ≥ 80, or current use of antihypertensive medication. Diabetes was defined by questionnaires where participants were asked if they had ever been diagnosed with diabetes by a healthcare provider. Use of antihypertensive and lipid-lowering treatments was collected through interviewer-administered questionnaires. Weight and height were measured with participants wearing light clothing and no shoes. Body weight was measured to the nearest 0.2 kg on a calibrated scale, whereas height was measured to the nearest 0.5 cm using a fixed vertical ruler. BMI was calculated as weight in kilograms divided by height in meters squared. Based on interviewer-administered questionnaires, cigarette smoking was classified as never, former, or current at each CARDIA visit. The CARDIA Physical Activity History questionnaire was used to estimate weekly leisure, occupational, and household physical exertion over the past 12 months.^31^

#### Statistical Analysis

To describe the study population, means and standard deviations or medians and interquartile ranges were used for continuous variables, and percentages were used for dichotomous variables. To test differences between groups, we used T-Test for continuous variables and Fisher’s exact test for categorical variables. To evaluate which variables were statistically significantly associated with each cognitive test, we used linear regression adjusting for age and education (Supplemental Table 1). For longitudinal analyses, we calculated the difference in cognitive test score (Visit 30 – Visit 25). To test the longitudinal association of IMAT with 5-year change in cognition, we used multiple linear regression, including adjustment for age, sex, race, education, study center, muscle volume, baseline cognitive test score, time of day, smoking, physical activity, diabetes, hypertension, and lipid-lowering medication use. We performed sensitivity analyses by also adjusting for BMI. We tested that all assumptions for linear regression models were met and found no violations, suggesting that there is no significant collinearity in our modeling procedure.

We tested an interaction term for both sex and race with IMAT, and neither was statistically significant (p-value=0.4233 and p-value= 0.7442, respectively); in supplemental tables, we present sex-stratified analyses. We present our results in terms of 1 Standard Deviation in IMAT to enhance the reliability and interpretability of our regression models. All statistical analyses were conducted using STATA version 18 (StataCorp, College Station, TX, USA) with significance at α=0.05.

## RESULTS

Among participants included in these analyses, 57% were women, and 46% were Black participants with a mean age of 50 2 ± 3.6 at Y25 (Table 1). Compared to White participants, Black participants were more likely to have lower DSST and RAVLT scores and higher Stroop scores (each indicating worse performance) than White Participants. Black participants were also more likely to have higher BMI, be hypertensive, have diabetes, and be on lipid-lowering medications. Black participants were also more likely to be current smokers and be less physically active. In this study, 95% of the participants had at least a high school education. Bivariate analyses were conducted with each of the study variables to inform our modeling strategies and to see which variables were related to DSST, RAVLT and Stroop (data not shown) in addition to known associations based on literature.

**Table 1.** Characteristics of Y25 CARDIA Cohort by Race.

|  | **Total**<br>**(N=2,512)** | **Black**<br>**(N=1,154)** | **White**<br>**(N=1,358)** | **p-value** |
| --- | --- | --- | --- | --- |
| **Age (years)** | 50.2 ± 3.6 | 49.6 ± 3.8 | 50.7 ± 3.3 | **< 0.001** |
| **Age Range** | 42-57 | 42-57 | 42-57 |  |
| **Female** | 1,432 (57.0%) | 701 (60.8%) | 731 (53.8%) | **< 0.001** |
| **Race (Black)** | 1,154 (46.0%) |  |  | **< 0.001** |
| **Study Center** |  |  |  | **< 0.001** |
| **Birmingham, AL** | 592 (23.6%) | 332 (28.7%) | 260 (19.2%) |  |
| **Chicago, IL** | 568 (22.6%) | 255 (22.1%) | 313 (23.1%) |  |
| **Minneapolis, MN** | 634 (25.2%) | 210 (18.2%) | 424 (31.2%) |  |
| **Oakland, CA** | 718 (28.6%) | 257 (30.9%) | 361 (26.6%) |  |
| **DSST Score** | 70.7 ± 15.7 | 64.9 ± 15.4 | 75.5 ± 14.4 | **< 0.001** |
| **DSST Score Percent Change** | -2.8 ± 21.8 | -3.4 ± 28.9 | -2.2 ± 12.7 | **0.177** |
| **RAVLT Score** | 9.1 ± 1.9 | 8.4 ± 1.8 | 9.6 ± 1.8 | **< 0.001** |
| **RAVLT Score Percent Change** | 2.8 ± 17.5 | 2.1 ± 19.0 | 3.4 ± 16.2 | 0.067 |
| **Stroop Score** | 22.5 ± 10.5 | 26.7 ± 11.9 | 19.2 ± 7.8 | **< 0.001** |
| **Stroop Score Percent Change** | 6.5 ± 49.5 | 7.9 ± 60.5 | 5.4 ± 38.3 | 0.223 |
| **Intermuscular Fat Volume (cm<sup>3</sup>)** | 2.3 ± 1.6 | 2.4 ± 1.7 | 2.2 ± 1.5 | **0.016** |
| **Muscle Volume (cm<sup>2</sup>)** | 20.3 ± 5.2 | 21.0 ± 5.4 | 19.7 ± 5.0 | **< 0.001** |
| **Waist Circumference (cm)** | 94.1 ± 15.6 | 96.7 ± 15.3 | 91.8 ± 15.5 | **< 0.001** |
| **BMI (kg/m<sup>2</sup>)** | 30.1 ± 7.0 | 31.9 ± 7.4 | 28.4 ± 6.2 | **< 0.001** |
| **Hypertension (yes)** | 1,175 (46.8%) | 729 (63.2%) | 446 (32.8%) | **< 0.001** |
| **Diabetes (yes)** | 261 (10.4%) | 163 (14.1%) | 98 (7.2%) | **< 0.001** |
| **Lipid Lowering Medications (yes)** | 382 (15.2%) | 192 (16.6%) | 190 (14.0%) | 0.086 |
| **Total years of Education** | 15.1 ± 2.6 | 14.2 ± 2.4 | 15.9 ± 2.5 | **< 0.001** |
| **Education (yes) (≥ high school)** | 2,391 (95.2%) | 1,059 (91.8%) | 1,332 (98.1%) | **< 0.001** |
| Smoke (yes) | 380 (15.1%) | 239 (20.7%) | 141 (10.4%) | < **0.001** |
| Physical Activity (points) | 338.5 ± 275.6 | 286.7 ± 266.9 | 382.5 ± 275.2 | < **0.001** |

In Table 2, we present the results for the association of IMAT at Year 25 with a 5-year change in DSST score. We found that one SD higher in IMAT was associated with a 0.57-point decrease in DSST score (p-value=0.018) in the minimally adjusted model (age, sex, race, education, center, muscle volume,Y25 DSST Score, and time of day) and a 0.52-point decrease in the model additionally adjusted for smoking and physical activity (p=0.035). After further adjustments for diabetes, hypertension, lipid-lowering medication (Model 3), this association was attenuated (β: -0.45, p=0.067), and when further adjusting for BMI (Table S3: Model 4) the association was lost (β: -0.12, p=0.667). We also tested for an association of IMAT with a change in DSST score stratified by race. There were no statistically significant findings between IMAT and 5-year change in DSST among Black participants. In contrast, among White participants, we found that one SD higher IMAT was associated with a 0.73-point decrease in DSST score in Model 1 (p-value=0.040)and in Model 2 (β:-0.73, p-value=0.044). The association was attenuated after adjusting for diabetes, hypertension, lipid-lowering medication (Model 3; β: -0.64, p=0.084), and overall adiposity (BMI; Table 3S Model 4; β: -0.16, p=0.704). Stratifying by sex, in Black women, one SD higher IMAT was associated with a 0.97-point decrease in DSST score (p-value=0.049) after adjusting for age, education, center, muscle volume, Y25 DSST Score, time of day, lifestyle and cardiometabolic diseases (Table 2S). In sensitivity analysis, when adjusting for BMI, the association was no longer significant (Table 3S).

**Table 2.** Association of IMAT with 5-year Change in DSST score in the CARDIA Cohort.

|  | Model 1 | Model 2 | Model 3 |
| --- | --- | --- | --- |
| **Total Sample**<br>(n=2,512) | -0.57 (0.241)<br>**0.018** | -0.52 (0.247)<br>**0.035** | -0.45 (0.248)<br>0.067 |
| **Race** |  |  |  |
| **Black Participants**<br>(n=1,154) | -0.55 (0.334)<br>0.099 | -0.44 (0.339)<br>0.195 | -0.42 (0.341)<br>0.216 |
| **White Participants**<br>(n=1,358) | -0.73 (0.354)<br>**0.040** | -0.73 (0.364)<br>**0.044** | -0.64 (0.367)<br>0.084 |

\*Results presented as beta coefficients (standard error) and p-value.
**Bold indicates** $P < 0.05$ .
Model 1: age, sex, race, education, center, and muscle volume, DSST Score from visit 25, and time
Model 2: M1 + smoke, physical activity
Model 3: M2 + diabetes, hypertension, lipid-lowering medication

**Table 3.**
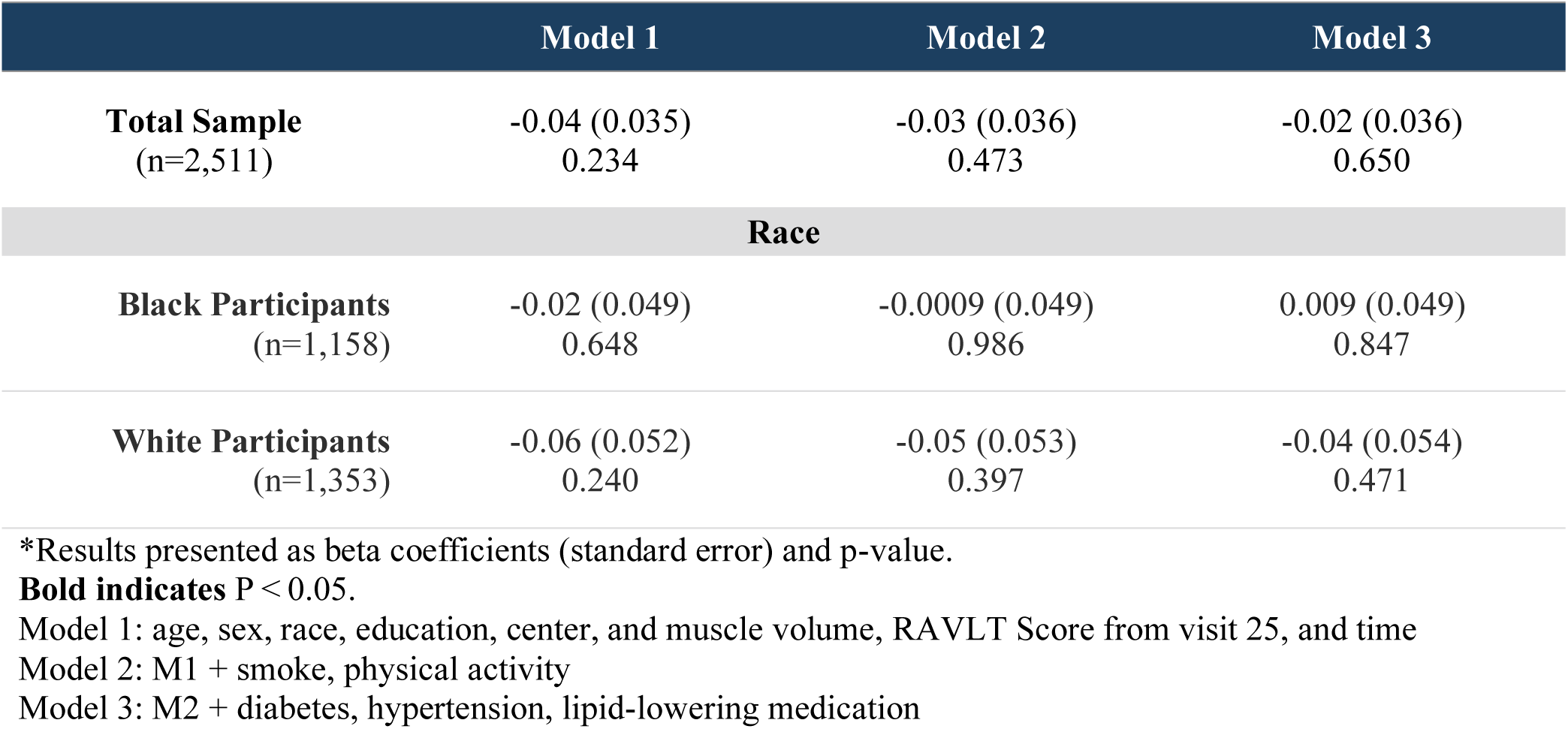
Association of IMAT with 5-year Change in RAVLT score in the CARDIA Cohort.

|  | Model 1 | Model 2 | Model 3 |
| --- | --- | --- | --- |
| **Total Sample**<br>(n=2,511) | -0.04 (0.035)<br>0.234 | -0.03 (0.036)<br>0.473 | -0.02 (0.036)<br>0.650 |
| **Race** |  |  |  |
| **Black Participants**<br>(n=1,158) | -0.02 (0.049)<br>0.648 | -0.0009 (0.049)<br>0.986 | 0.009 (0.049)<br>0.847 |
| **White Participants**<br>(n=1,353) | -0.06 (0.052)<br>0.240 | -0.05 (0.053)<br>0.397 | -0.04 (0.054)<br>0.471 |

\*Results presented as beta coefficients (standard error) and p-value.
**Bold indicates** $P < 0.05$ .
Model 1: age, sex, race, education, center, and muscle volume, RAVLT Score from visit 25, and time
Model 2: M1 + smoke, physical activity
Model 3: M2 + diabetes, hypertension, lipid-lowering medication

**Table 4.** Association of IMAT with 5-year Change in Stroop score in the CARDIA Cohort.

|  | Model 1 | Model 2 | Model 3 |
| --- | --- | --- | --- |
| **Total Sample**<br>(n=2,458) | 0.33 (0.230)<br>0.155 | 0.27 (0.235)<br>0.259 | 0.22 (0.237)<br>0.363 |
| **Race** |  |  |  |
| **Black Participants**<br>(n=1,111) | 0.24 (0.385)<br>0.531 | 0.09 (0.391)<br>0.820 | 0.07 (0.393)<br>0.868 |
| **White Participants**<br>(n=1,347) | 0.41 (0.265)<br>0.121 | 0.40 (0.273)<br>0.143 | 0.35 (0.275)<br>0.200 |

\*Results presented as beta coefficients (standard error) and p-value.
**Bold indicates** $P < 0.05$ .
Model 1: age, sex, race, education, center, and muscle volume, Stroop Score from visit 25, and time
Model 2: M1 + smoke, physical activity
Model 3: M2 + diabetes, hypertension, lipid-lowering medication

There were no statistically significant findings between IMAT and 5-year change in RAVLT.

When assessing the association between IMAT and 5-year change in Stroop, we found no significant associations in the total sample or by race group. In Supplemental Table 2 we show that one SD higher in IMAT was associated with a 1.02 point increase in Stroop score for all women (p-value=0.003) and 1.05 points higher specifically among Black women (p-value=0.043), after adjusting for age, education, center, muscle volume, Y25 Stroop score, time of day, smoking, physical activity, diabetes, hypertension, and lipid-lowering medication. In White women (Table 2S), we saw a significant association in Model 2 and 3 (β:0.92, p=0.027 and β:0.91, p=0.030, respectively).

Supplemental tables S4-S6 present the cross-sectional results for the association of IMAT with each of the cognitive tests. In the total sample, we found a statistically significant association between IMAT and each of the administered tests, specifically in women and in White participants, thus confirming the findings of the previous studies.

## DISCUSSION

In this community-based longitudinal study of Black and White middle-aged men and women, we found that CT-derived abdominal myosteatosis was associated with a decline in performance on the DSST, an early indicator of future dementia risk. In race-stratified analyses, we found these associations were significant for White but not Black participants. Demographic and other risk factor characteristics did not explain these associations, however, they were attenuated after adjustment for diabetes, hypertension, and lipid-lowering medication. We found no significant associations for IMAT with the Stroop or RAVLT in the full sample or stratified by race. Our findings suggest that myosteatosis may affect neurocognitive domains differently, warranting further research to better understand this relationship, especially in the context of cardiometabolic diseases.

Research on the relationship between myosteatosis and cognitive function is gaining momentum. Although the exact nature of this relationship is still under investigation, some studies have provided some insights into their potential associations. Previous studies have linked myosteatosis with lower psychomotor function and visual learning^21^ and older-appearing brain and increased rate of brain aging in brain MRI studies.^22^ Our findings support and contribute to this body of evidence. There is an increasing awareness that general and central adiposity and loss of muscle mass are risk factors for future dementia risk, and our results, combined with other recent research, point toward skeletal muscle adiposity as an additional risk factor for cognitive decline. Our findings are consistent with results from a cross-sectional study that found that greater calf skeletal muscle density was associated with better performance in the DSST and thus better information processing speed among middle-aged (55–65 years) women, independent of relevant covariates.^20^ Another among very few studies focusing on myosteatosis and cognition is a recent longitudinal study cohort of older Black and White adults. This study found a 5-year increase in thigh IMAT was associated with a 5-year decline in mini-mental state exam (3MS)^23^, independent of overall adiposity, muscle health, and demographic factors.^23^ Although we did not find a significant association between greater IMAT at Year 25 with a decline in DSST score in Black participants in comparison to this study, the percent change in DSST score was greater in Black (-3.4%) than in White participants (-2.2%). These results were unexpected, as skeletal muscle adiposity is higher and increases more with age in Black individuals compared with White individuals, independent of general and central adiposity.^32,33^ The reason for these unexpected findings in our study is not clear, but it is possible that greater myosteatosis in middle age may have a greater effect on cognitive impairment in White individuals while for Black individuals these effects may occur later in life. Future studies should include a wide age range of middle-aged and older multi-ethnic populations.

### Biological Plausibility

Ectopic adipose tissue infiltration in skeletal muscle can impair the normal physiologic function of the organ, making it a pathophysiologically relevant fat depot that may play a key role in the risk of metabolic abnormalities such as insulin resistance and type 2 diabetes.^16,17,19^ Myosteatosis increases inflammation^16^, which has also been linked to brain health and healthy aging.^34^ A possible mechanism for this relationship is through higher glucose and lower insulin sensitivity, which have a detrimental effect on brain parenchyma.^35^ Although myosteatosis is associated with cardio-metabolic disorders, and these are plausible intermediaries for the association between myosteatosis and poor cognition, adjustment for these and other traditional risk factors didn’t explain the observed associations in this study. Another pathway may be the impairment of the endocrine function of the muscle. Due to proximity to the muscle fiber, muscle adiposity may impact local muscle metabolism as well as systemic metabolism by impairment of myokine secretion and/or by increased production of pro-inflammatory adipokines and with increased macrophage invasion in the muscle.^36,37^ Mitochondrial dysfunction has been previously linked to greater myosteatosis^38^ and could also play a role in cognitive decline for individuals with higher amounts of myosteatosis. Thus, in addition to impaired glucose and insulin resistance, increased secretion of pro-inflammatory myokines and adipokines and poor mitochondrial function could be additional mechanisms underlying an association of greater myosteatosis and cognitive decline, but further study is needed.

Evidence of a connection between myosteatosis, body mass index (BMI), and other forms of ectopic fat deposits is not very strong.^39^ The relationship of other types of ectopic adiposity depots, such as visceral adiposity, with brain health^12,40^ has been extensively studied, yet the relationship between ectopic skeletal muscle adiposity with brain health and cognitive function is still in the early stages of investigation. For example, the degree of association between various adipose depots and cardiometabolic disease varies by depot type,^14,15^ suggesting that the potential adverse contribution of ectopic adiposity on brain health might also differ. There may be factors specific to the accumulation of adiposity within skeletal muscles that vary from other ectopic adiposity storage sites. Abdominal muscles are largely utilized in stabilization of the thorax, while thigh muscles are used principally for locomotion. These differences can have effects on energy metabolism by muscle group and could explain some of our findings and discrepancies with previous studies. It also implies that the health consequences of ectopic fat deposits may vary depending on where the excess fat is located.

When adjusting for baseline body mass index (i.e., a marker of overall adiposity) the associations between myosteatosis and cognitive function decline were no longer significant. This could be because results are not adipose tissue depot-specific. The amount of adipose tissue in various depots may be directly linked to cognitive function, which might clarify why the association attenuates when we adjust for BMI. It is also possible that both cognitive impairment and greater myosteatosis contribute to overall adiposity. In this case, adjusting for total adiposity (i.e., BMI) would be adjusting for a collider and thus, an over-adjustment bias.

This study has potential limitations. We only assessed Black and White individuals; thus, results are not generalizable to other racial and ethnic groups. A smaller sample size of Black men might have precluded the subgroup examination. Future studies should focus on a large population-based sample of Black men and/or studies that oversample Black men. It is possible that Black men were less likely to attend follow-up visits than White men due to life circumstances, and competing risks (survival bias), among other reasons.^41,42^ Another important limitation is that CT-derived measures were assessed only once in the abdomen. Repeated measures of myosteatosis can enhance our understanding of the relationship with cognitive decline and facilitate analyses to examine changes in adiposity related to cognitive decline. Due to the lack of data on myokines and adipokines and mitochondrial function, we were unable to explore potential mechanisms that may link myosteatosis with poor cognition. Although we found significant findings for abdominal IMAT and a 5-year decline in DSST and Stroop score in women and White participants, the RAVLT was not significantly associated with cognitive decline. It is unclear whether or why such relationships would vary across different cognitive domains. Future studies should look at a larger battery of cognitive tests, different locations of myosteatosis (abdomen and extremities) with longer follow-ups, and various racial groups to better understand this association and the possible biological mechanisms behind it. Regardless, this study has very notable strengths, including a large sample size of middle-aged men and women from two racial groups. We have objective CT measures of myosteatosis and two timepoint measures of cognitive function. Additionally, we have extensive measures of potential confounding variables such as interviewer-administered questionnaires that assessed broad health, disease, sociodemographic, and lifestyle information.

## CONCLUSION

In conclusion, in this longitudinal study of a biracial middle-aged cohort, we showed that abdominal myosteatosis may be a novel risk factor for cognitive decline, especially in White adults. Next steps in this line of research include further longitudinal studies with a longer follow-up, in other populations across a wide age range, using myosteatosis from various muscle groups, as well as determining what biological mechanisms of action may be responsible for the link between myosteatosis and cognitive decline and dementia risk. Our findings also underscore the need for a better understanding of women’s reproductive health and the potential influence of menopausal stages. This may elucidate the difference in the association between myosteatosis and cognitive function in women compared to men.

## Data Availability

Data from CARDIA are available to investigators.

## ACKNOWLEDGMENTS

The authors thank all CARDIA participants for their commitment to the study and the many CARDIA investigators who have contributed to its innovative science through the years.

## DATA AVAILABILITY

Data from CARDIA are available to investigators.

---

**Table S1.** Characteristics of CARDIA Cohort by Completion of DSST by visits.

|  | Completed DSST at V25<br>and V30<br>N=(2,512)<br>(Analytical Sample) | Did NOT complete DSST<br>at V25 and V30<br>(N=614) | p-value |
| --- | --- | --- | --- |
| Age (years) | 50.1 ± 3.6 | 50.0 ± 3.8 | 0.432 |
| Age range | 42-57 | 42-56 | - |
| Sex (female) | 1,423 (56.9%) | 990 (50.4%) | < **0.001** |
| Race (Black) | 1,351 (54.0%) | 829 (42.2%) | < **0.001** |
| DSST Score | 70.7 ± 15.7 | 66.6 ± 16.8 | < **0.001** |
| Total Muscle Density (HU) | 35.1 ± 3.0 | 35.1 ± 2.7 | 0.812 |
| Visceral Adipose Tissue (cm<sup>3</sup>) | 130.6 ± 73.5 | 137.9 ± 75.3 | **0.045** |
| Intermuscular Fat Volume (cm<sup>3</sup>) | 4.8 ± 0.6 | 4.9 ± 0.6 | **0.009** |
| Muscle Volume (cm<sup>2</sup>) | 246.5 ± 60.6 | 256.8 ± 69.2 | **0.002** |
| Height (cm) | 170.3 ± 9.4 | 170.2 ± 9.6 | 0.840 |
| Waist Circumference (cm) | 94.1 ± 15.7 | 95.7 ± 16.9 | **0.024** |
| BMI (kg/m<sup>2</sup>) | 30.1 ± 7.0 | 30.7 ± 7.5 | 0.053 |
| Systolic BP (mmHg) | 119.1 ± 15.7 | 122.1 ± 17.7 | < **0.001** |
| Diastolic BP (mmHg) | 74.5 ± 11.2 | 76.4 ± 11.4 | < **0.001** |
| Hypertension (yes) | 1,173 (46.9%) | 1,690 (86.1%) | < **0.001** |
| Type 2 Diabetes (yes) | 261 (10.4%) | 82 (13.5%) | **0.004** |
| Lipid Lowering Medications (yes) | 381 (15.2%) | 101 (16.6%) | 0.190 |
| Years of education | 15.1 ± 2.7 | 14.8 ± 2.6 | **0.002** |
| Education (yes )(≥high-school) | 2,381 (95.2%) | 1,926 (98.0%) | <**0.001** |
| Current Smoker (yes) | 378 (15.1%) | 145 (23.9%) | <**0.001** |
| Physical Activity (points) | 338.1 ± 275.7 | 339.1 ± 274.2 | 0.936 |

**Table S2.** Association of IMAT with 5-year Change in Cognitive Tests in the CARDIA Cohort by Sex and Race.

|  | Model 1 | Model 2 | Model 3 |
| --- | --- | --- | --- |
| **DSST** |  |  |  |
| **Total Men**<br>(n=1,080) | -0.33 (0.300)<br>0.265 | -0.34 (0.309)<br>0.274 | -0.25 (0.312)<br>0.428 |
| **Black Men**<br>(n=453) | 0.02 (0.456)<br>0.958 | 0.12 (0.471)<br>0.794 | 0.11 (0.479)<br>0.824 |
| **White Men**<br>(n=627) | -0.74 (0.402)<br>0.066 | -0.84 (0.412)<br>**0.041** | -0.70 (0.414)<br>0.093 |
| **Total Women**<br>(n=1,432) | -0.88 (0.383)<br>**0.022** | -0.79 (0.389)<br>**0.042** | -0.76 (0.390)<br>0.052 |
| **Black Women**<br>(n=701) | -1.06 (0.489)<br>**0.031** | -0.98 (0.492)<br>**0.046** | -0.97 (0.491)<br>**0.049** |
| **White Women**<br>(n=731) | -0.66 (0.609)<br>0.278 | -0.56 (0.629)<br>0.377 | -0.51 (0.635)<br>0.420 |
| **RAVLT** |  |  |  |
| **Total Men**<br>(n=1,080) | -0.06 (0.050)<br>0.209 | -0.04 (0.051)<br>0.394 | -0.04 (0.052)<br>0.487 |
| **Black Men**<br>(n=457) | -0.04 (0.071)<br>0.590 | -0.01 (0.073)<br>0.896 | -0.004 (0.074)<br>0.952 |
| **White Men**<br>(n=623) | -0.08 (0.071)<br>0.286 | -0.06 (0.073)<br>0.384 | -0.05 (0.074)<br>0.517 |
| **Total Women**<br>(n=1,431) | 0.006 (0.052)<br>0.931 | 0.01 (0.052)<br>0.777 | 0.02 (0.053)<br>0.676 |
| **Black Women**<br>(n=701) | 0.02 (0.069)<br>0.741 | 0.03 (0.069)<br>0.641 | 0.04 (0.069)<br>0.595 |
| **White Women**<br>(n=730) | -0.03 (0.079)<br>0.693 | -0.009 (0.081)<br>0.912 | -0.01 (0.082)<br>0.893 |
| **Stroop** |  |  |  |
| **Total Men**<br>(n=1,052) | -0.27 (0.330)<br>0.417 | -0.49 (0.338)<br>0.150 | -0.53 (0.341)<br>0.118 |
| **Black Men**<br>(n=433) | -0.77 (0.598)<br>0.198 | -1.06 (0.613)<br>0.084 | -1.10 (0.621)<br>0.077 |
| **White Men**<br>(n=619) | 0.16 (0.360)<br>0.657 | -0.04 (0.368)<br>0.909 | -0.15 (0.371)<br>0.688 |
| **Total Women**<br>(n=1,406) | 0.98 (0.331)<br>**0.003** | 1.05 (0.336)<br>**0.002** | 1.02 (0.337)<br>**0.003** |
| **Black Women**<br>(n=678) | 1.12 (0.516)<br>**0.030** | 1.07 (0.520)<br>**0.040** | 1.05 (0.519)<br>**0.043** |
| **White Women**<br>(n=728) | 0.70 (0.402)<br>0.080 | 0.92 (0.414)<br>**0.027** | 0.91 (0.418)<br>**0.030** |

\*Results presented as beta coefficients (standard error) and p-value.
**Bold indicates** P < 0.05.
Model 1: age, sex, race, education, center, and muscle volume, DSST/RAVLT/Stroop Score from visit 25, and time
Model 2: M1 + smoke, physical activity
Model 3: M2 + diabetes, hypertension, lipid-lowering medication

**Table S3.** Sensitivity Analysis for the Association of IMAT with 5-year Change in DSST score in the CARDIA Cohort.

|  | Model 1 | Model 2 | Model 3 | Model 4 |
| --- | --- | --- | --- | --- |
| **Total Sample**<br>(n=2,512) | -0.57 (0.241)<br>**0.018** | -0.52 (0.247)<br>**0.035** | -0.45 (0.248)<br>0.067 | -0.12 (0.276)<br>0.667 |
| **Race** |  |  |  |  |
| **Black Participants**<br>(n=1,154) | -0.55 (0.334)<br>0.099 | -0.44 (0.339)<br>0.195 | -0.42 (0.341)<br>0.216 | -0.11 (0.369)<br>0.768 |
| **White Participants**<br>(n=1,358) | -0.73 (0.354)<br>**0.040** | -0.73 (0.364)<br>**0.044** | -0.64 (0.367)<br>0.084 | -0.16 (0.423)<br>0.704 |
| **Sex** |  |  |  |  |
| **Total Men**<br>(n=1,080) | -0.33 (0.300)<br>0.265 | -0.34 (0.309)<br>0.274 | -0.25 (0.312)<br>0.428 | 0.09 (0.350)<br>0.803 |
| **Black Men**<br>(n=453) | 0.02 (0.456)<br>0.958 | 0.12 (0.471)<br>0.794 | 0.11 (0.479)<br>0.824 | 0.40 (0.532)<br>0.448 |
| **White Men**<br>(n=627) | -0.74 (0.402)<br>0.066 | -0.84 (0.412)<br>**0.041** | -0.70 (0.414)<br>0.093 | -0.17 (0.471)<br>0.712 |
| **Total Women**<br>(n=1,432) | -0.88 (0.383)<br>**0.022** | -0.79 (0.389)<br>**0.042** | -0.76 (0.390)<br>0.052 | -0.40 (0.428)<br>0.349 |
| **Black Women**<br>(n=701) | -1.06 (0.489)<br>**0.031** | -0.98 (0.492)<br>**0.046** | -0.97 (0.491)<br>**0.049** | -0.66 (0.519)<br>0.206 |
| **White Women**<br>(n=731) | -0.66 (0.609)<br>0.278 | -0.56 (0.629)<br>0.377 | -0.51 (0.635)<br>0.420 | -0.07 (0.749)<br>0.926 |

\*Results presented as beta coefficients (standard error) and p-value.
**Bold indicates** P < 0.05.
Model 1: age, sex, race, education, center, and muscle volume, DSST Score from visit 25, and time
Model 2: M1 + smoke, physical activity
Model 3: M2 + diabetes, hypertension, lipid-lowering medication
Model 4: M3 + BMI

**Table S4.** Cross-sectional Association of IMAT with DSST in CARDIA Cohort.

|  | Model 1 | Model 2 | Model 3 |
| --- | --- | --- | --- |
| **Total Sample**<br>(n=2,512) | -1.53 (0.360)<br>**< 0.001** | -1.15 (0.366)<br>**0.002** | -1.07 (0.367)<br>**0.004** |
| **Race** |  |  |  |
| **Black Participants**<br>(n=1,154) | -1.56 (0.513)<br>**0.002** | -1.09 (0.517)<br>**0.036** | -1.05 (0.518)<br>**0.042** |
| **White Participants**<br>(n=1,358) | -1.58 (0.517)<br>**0.002** | -1.27 (0.529)<br>**0.017** | -1.17 (0.534)<br>**0.028** |
| **Sex** |  |  |  |
| **Total Men**<br>(n=1,080) | -0.61 (0.501)<br>0.220 | -0.20 (0.511)<br>0.688 | -0.07 (0.514)<br>0.891 |
| **Black Men**<br>(n=453) | -0.18 (0.733)<br>0.811 | 0.34 (0.749)<br>0.648 | 0.42 (0.759)<br>0.584 |
| **White Men**<br>(n=627) | -1.08 (0.690)<br>0.118 | -0.78 (0.704)<br>0.268 | -0.64 (0.710)<br>0.368 |
| **Total Women**<br>(n=1,432) | -2.30 (0.535)<br>**< 0.001** | -1.99 (0.538)<br>**< 0.001** | -1.95 (0.540)<br>**< 0.001** |
| **Black Women**<br>(n=694) | -2.58 (0.733)<br>**< 0.001** | -2.23 (0.727)<br>**0.002** | -2.20 (0.726)<br>**0.003** |
| **White Women**<br>(n=731) | -1.96 (0.792)<br>**0.014** | -1.64 (0.816)<br>**0.045** | -1.63 (0.825)<br>**0.048** |

\*Results presented as beta coefficients (standard error) and p-value.
**Bold indicates** P < 0.05.
Model 1: age, sex, race, education, center, and muscle volume, Stroop Score from visit 25, and time
Model 2: M1 + smoke, physical activity
Model 3: M2 + diabetes, hypertension, lipid-lowering medication

**Table S5.** Cross-sectional Association of IMAT with RAVLT in CARDIA Cohort.

|  | Model 1 | Model 2 | Model 3 |
| --- | --- | --- | --- |
| **Total Sample**<br>(n=2,511) | -0.15 (0.044)<br>**0.001** | -0.15 (0.045)<br>**0.001** | -0.14 (0.045)<br>**0.002** |
| **Race** |  |  |  |
| **Black Participants**<br>(n=1,158) | -0.13 (0.060)<br>**0.038** | -0.09 (0.061)<br>0.149 | -0.09 (0.061)<br>0.162 |
| **White Participants**<br>(n=1,353) | -0.16 (0.065)<br>**0.015** | -0.20 (0.066)<br>**0.003** | -0.17 (0.067)<br>**0.010** |
| **Sex** |  |  |  |
| **Total Men**<br>(n=1,080) | -0.11 (0.063)<br>0.081 | -0.12 (0.065)<br>0.067 | -0.10 (0.066)<br>0.117 |
| **Black Men**<br>(n=457) | -0.006 (0.091)<br>0.947 | 0.006 (0.094)<br>0.950 | 0.01 (0.095)<br>0.896 |
| **White Men**<br>(n=623) | -0.19 (0.088)<br>**0.031** | -0.22 (0.090)<br>**0.015** | -0.20 (0.091)<br>**0.026** |
| **Total Women**<br>(n=1,431) | -0.19 (0.063)<br>**0.003** | -0.18 (0.064)<br>**0.004** | -0.18 (0.064)<br>**0.006** |
| **Black Women**<br>(n=701) | -0.22 (0.084)<br>**0.009** | -0.18 (0.083)<br>**0.032** | -0.18 (0.083)<br>**0.035** |
| **White Women**<br>(n=731) | -0.15 (0.098)<br>0.130 | -0.21 (0.101)<br>**0.038** | -0.19 (0.101)<br>0.063 |

\*Results presented as beta coefficients (standard error) and p-value.
**Bold indicates** P < 0.05.
Model 1: age, sex, race, education, center, and muscle volume, RAVLT Score from visit 25, and time
Model 2: M1 + smoke, physical activity
Model 3: M2 + diabetes, hypertension, lipid-lowering medication

**Table S6.**
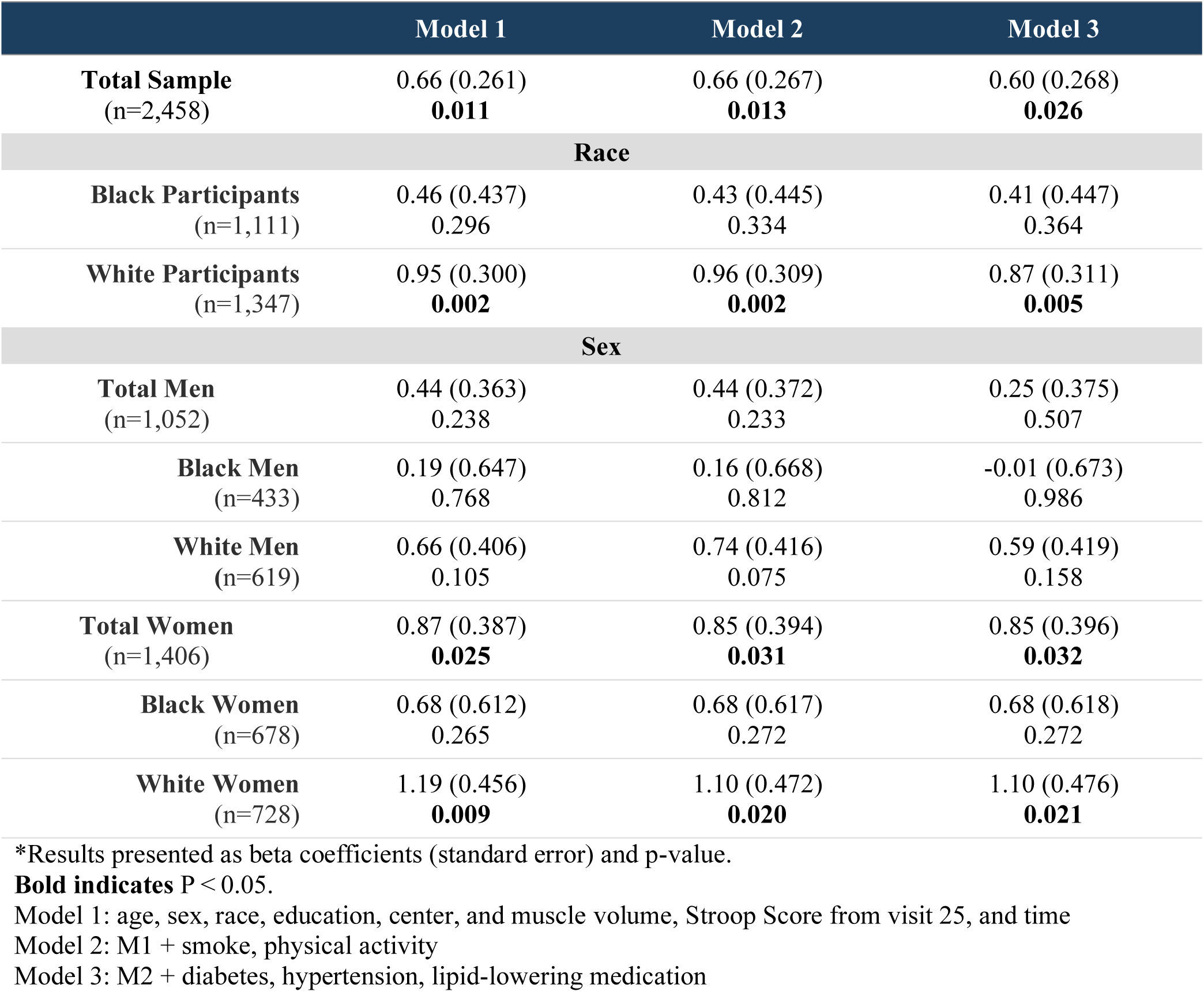
Cross-sectional Association of IMAT with Stroop in CARDIA Cohort.

|  | Model 1 | Model 2 | Model 3 |
| --- | --- | --- | --- |
| **Total Sample**<br>(n=2,458) | 0.66 (0.261)<br>**0.011** | 0.66 (0.267)<br>**0.013** | 0.60 (0.268)<br>**0.026** |
| **Race** |  |  |  |
| **Black Participants**<br>(n=1,111) | 0.46 (0.437)<br>0.296 | 0.43 (0.445)<br>0.334 | 0.41 (0.447)<br>0.364 |
| **White Participants**<br>(n=1,347) | 0.95 (0.300)<br>**0.002** | 0.96 (0.309)<br>**0.002** | 0.87 (0.311)<br>**0.005** |
| **Sex** |  |  |  |
| **Total Men**<br>(n=1,052) | 0.44 (0.363)<br>0.238 | 0.44 (0.372)<br>0.233 | 0.25 (0.375)<br>0.507 |
| **Black Men**<br>(n=433) | 0.19 (0.647)<br>0.768 | 0.16 (0.668)<br>0.812 | -0.01 (0.673)<br>0.986 |
| **White Men**<br>(n=619) | 0.66 (0.406)<br>0.105 | 0.74 (0.416)<br>0.075 | 0.59 (0.419)<br>0.158 |
| **Total Women**<br>(n=1,406) | 0.87 (0.387)<br>**0.025** | 0.85 (0.394)<br>**0.031** | 0.85 (0.396)<br>**0.032** |
| **Black Women**<br>(n=678) | 0.68 (0.612)<br>0.265 | 0.68 (0.617)<br>0.272 | 0.68 (0.618)<br>0.272 |
| **White Women**<br>(n=728) | 1.19 (0.456)<br>**0.009** | 1.10 (0.472)<br>**0.020** | 1.10 (0.476)<br>**0.021** |

\*Results presented as beta coefficients (standard error) and p-value.
**Bold indicates** $P < 0.05$ .
Model 1: age, sex, race, education, center, and muscle volume, Stroop Score from visit 25, and time
Model 2: M1 + smoke, physical activity
Model 3: M2 + diabetes, hypertension, lipid-lowering medication

